# The role of healthcare cost accounting in pricing and reimbursement in low- and middle-income countries

**DOI:** 10.1101/2022.05.19.22275296

**Authors:** Lorna Guinness, Srobana Ghosh, Abha Mehndiratta, Hiral Anil Shah

**Affiliations:** Center for Global Development, London, United Kingdom; Center for Global Infectious Disease Analysis, Department of Infectious Disease Epidemiology, School of Public Health, Imperial College London, London, UK

**Author notes:** <u>Corresponding author</u> Lorna Guinness, Great Peter House, Abbey Gardens, Great College St, London, SW1P 3SE.

## Abstract

**Background:** Progress towards universal health coverage requires evidence based policy and good quality cost data systems. Establishing these systems, can be complex, resource intensive and take time. Synthesising information on the experiences of low-and-middle income countries (LMICs) in trying to institutionalize cost data systems provides lessons for other settings.

**Methods:** We performed a scoping literature review, screening for publicly available peer-reviewed English-language publications alongside a grey literature search, focussing on national level health system costing in LMICs, and used a narrative synthesis approach to document experience of costing and how costing evidence is used to inform tariffs.

**Results:** A total of 484 papers were initially identified of which 30 papers were considered eligible. Fourteen papers reported on primary cost data collection for price-setting purposes; 18 papers provided an explanation of how cost evidence informs tariff-setting. Documented experience is largely focussed in the Asia region (n = 22) with countries at different stages of developing cost systems to inform tariff setting. Country experiences on healthcare cost accounting tend to showcase country costing experiences, methods and implementation. There is little documentation of how cost data has been incorporated into decision making and price setting. Where cost data, cost systems and costing has been used, improved transparency in decision making alongside increased service provision efficiency has followed.

**Conclusions:** While there are accepted and widely used methods for generating cost information, countries need to build sustainable cost systems appropriate to their settings and budgets and adopt transparent processes and methodologies for translating costs into prices.

## INTRODUCTION

Low-and-middle income countries (LMICs) have been making significant progress towards universal health coverage through innovative healthcare financing. One focus of healthcare financing reforms has been reimbursement schemes that target the explicit goals of efficiency and cost containment while improving quality and reaching the poor and vulnerable. Historically, block grants have been used to reimburse healthcare providers in publicly financed systems in LMICs. However, as national-level public purchasers have evolved and a broader range of healthcare providers (e.g. private or faith-based healthcare providers) are accepted as part of the developing health system, newer prospective payment mechanisms and systems of provider reimbursement are being used by government purchasers of healthcare [1].

Common prospective payment mechanisms such as case-based payments for the reimbursement of secondary and/or tertiary care and capitation payments for primary care providers are now being championed across developing regions and countries. Case-based payments are equivalent to a system where providers are reimbursed based on cases treated rather than per service or per bed days [2]. On the other hand, capitation based payments are equivalent to a payment system where lump-sum payments are made to care providers based on the number of patients in a target population [2,3].

Setting reimbursement rates requires a reliable cost evidence base to enable price negotiations that are transparent, facilitate cost control and help drive providers to more efficient services. In principle, information is needed on the average cost per case across all admissions and/or visits (a base rate) and the relative value of different conditions as classified in the respective country (e.g., Diagnosis Related Groups, specialty-based classification, intervention specific health benefit package etc) [4–6]. In a case-based payment scheme, the service groups are often DRGs or a similar grouping system which provides a means of relating the type of patients a hospital treats to the costs incurred by the hospital. For capitation-based systems the grouping is related to the average expected cost of treating a patient under the care of the provider. In both types of system, the technical process of price-setting requires a robust cost system to be in place, using principles that can be guided or even mandated by a purchaser, in order to generate reliable health service cost estimates.

Raulinajtys-Grzybek (2014) defines the cost system as “*a cost accounting system that ensure the cost homogeneity of individual groups* (of services)” [7]. There are however variation in costing systems across health systems as a result of choices about the process of collecting and verifying the data, the stage of development of the reimbursement system, the regulation around cost accounting and the costing methodology used [7]. For example, they can vary from one off costing studies to regular national costing surveillance [7,8]. Some cost surveys involve all participating providers e.g., in the UK and Australia all providers are mandated to submit cost accounting information; in others, only a sample of representative providers is used e.g. France, Germany and Thailand [9].

In terms of costing methodology, according to Gapenski and Reiter (2016) “the holy grail of cost estimation is costing at the service or individual patient level” [10]. More advanced systems e.g., those in the UK and Australia use bottom-up style costing methods to derive patient level DRG costs [11]; but there are simplified methods available that calculate the average cost of procedure through step down allocation methods [12]. Whichever approach is taken, it is important that the costing is nationally acceptable and can capture structural differences in cost that might be present (types of provider, demography, geography etc) as well as variability between the cost of the conditions treated. In addition, the national costing system should be standardised across providers, creating transparency and comparability [8].

In LMICs, while the process of payment reform has been well documented, there is less information available about the role of cost information in the technical process of setting reimbursement rates. Increasing number of countries are moving towards case-based payment schemes for secondary care within their UHC strategies. Documenting the cost systems used to generate evidence for rate setting can provide lessons for the further development of existing systems or the establishment of new ones. The aim of this paper is therefore to synthesise the evidence on the role of cost accounting in setting reimbursement rates for case-based payment schemes in LMICs. We perform a scoping literature review and narrative synthesis to document the current practice in LMICs based on publicly available information and recommend steps for the technical process of price-setting in LMICS in the context of UHC goals.

## METHODS

### Search Strategy and Selection Process

A scoping review approach was used to synthesise the evidence on cost accounting in LMICs. We aimed to map the body of literature, clarify key concepts and identify any gaps in the research [13]. We further refined our research question using a standard PICO framework:

- Problem: technical process for price setting for hospital case-based payments in LMICs
- Intervention: cost systems
- Comparator: non-cost-based methods
- Outcome: improved evidence base for decision-making

We used several approaches in identifying the literature. First, we conducted a search of the literature for peer-reviewed English-language publications indexed in Pubmed, Medline, Econlit and in the Web of Science on the subject of national level health system costing in LMICs and the associated design of their costing systems. Our search was conducted using the following terms: (“case*mix” or “cost systems” or “cost*accounting” or “ref*costs” or “resource weights” or “cost*weights” or “national reimbursement” or “DRG” or “hospital payment systems” or “fee*for*service”) AND (“LMIC” or “low resource settings” or “developing countries”). We conducted a search that included the country name of all LMICs, as defined by the World Bank. To complement this, we consulted existing libraries of both grey and peer-reviewed literature held by the research team. We then conducted an analysis of text words contained in the title and abstract to help identify further keywords and index terms. A further search was then conducted using the identified keywords and index terms. Finally, the reference list of all identified reports and articles were reviewed for any reports or papers that might have been missed. The search strategy is provided in the Supplement.

### Eligibility Criteria & Screening

Papers in the English language and published since 2000 were included. Results were then hand screened to ensure that the topic was limited to the eligible countries (LMICs as defined by the World Bank) and that the study identified and/or described the development of the national tariffs for hospital reimbursements and/or the methods used to estimate or inform the tariffs for hospital services reimbursement. The titles and abstracts were screened independently by two reviewers as per the inclusion and exclusion criteria defined by the study. The second screen involved reviewing full texts.

### Data Extraction & Synthesis

The papers were then classified according to whether they explained the technical process of price-setting for reimbursements (i.e., if and how cost data was used) and whether they reported on the process of primary cost data collection for price-setting. For those papers or case studies reporting on the process of primary cost data collection for price setting, we extracted information on the method of cost data collection, the output and any commentary on how the cost data was used for price setting for hospital case-based payments including identifying the commissioning agency. From the papers that described how cost data is used in price setting, we extracted information on any description of the technical aspects of the tariff setting system in place, at the time of the study, and the key strengths and challenges of the approach used. For those papers describing more than one country experience, only evidence on LMIC experience was extracted. We use a narrative review approach to summarise the evidence by country. Data extraction was performed by one reviewer and then checked independently.

## RESULTS

### Overview of the literature

A total of 484 papers were initially identified of which 424 papers were excluded in the initial screening. The second screen involved reviewing full texts, leading to the inclusion of 30 papers in the review as described in the PRISMA diagram in Figure 1.

**Figure 1:**
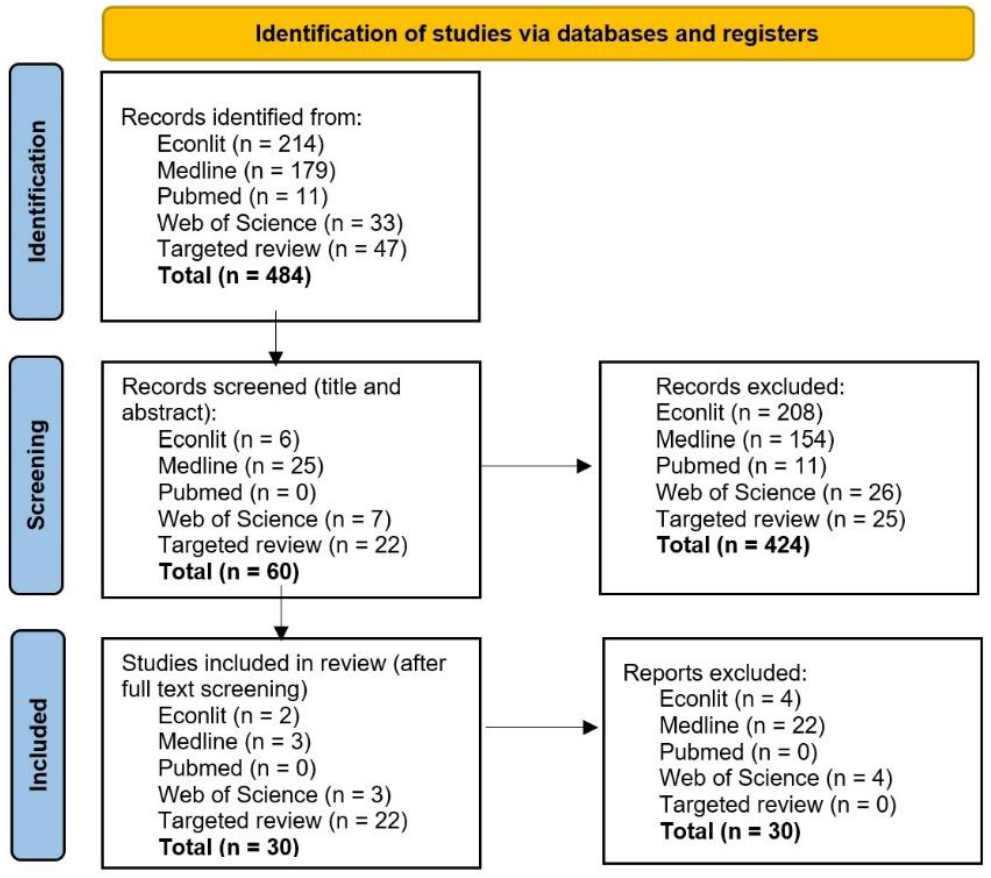
PRISMA diagram.

Of the 30 papers extracted, 7 papers stated a global focus (including LMICs) [1,4,14–18] and one paper reported to be focussed on the Asia region [19] (see Figure 2). Of the single country focussed papers, 6 related to India [20–25]. We found 3 studies each related to Thailand [6,26,27] and Vietnam [28–30]. There were 2 studies focussed on each of Indonesia [31,32], Iran [33,34], Malaysia [35,36] and Cambodia [37,38] and one study each for Kenya [39] and China [40]. Further, within the global papers, we identified case studies on: Kazakhstan, Kyrgyzstan, India, Malaysia, Thailand, and China.

**Figure 2:**
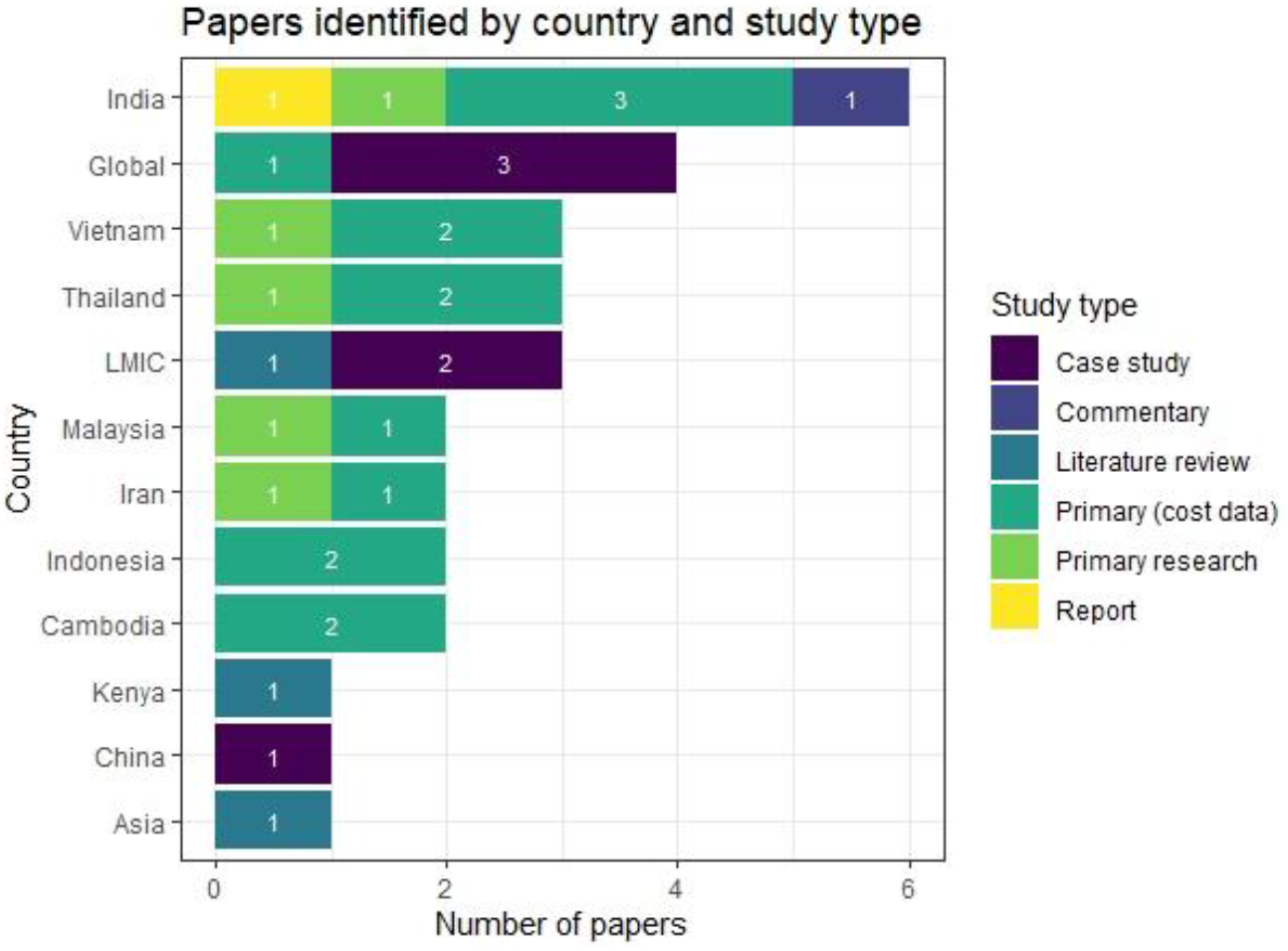
Number of papers by country breakdown and types of study.

### Papers reporting on primary collection of cost data to inform tariff-setting

Twenty-three case studies from 14 fourteen studies reported on primary cost data collection for price setting purposes in a single country setting, either describing methods or both methods and results (see Table 1). Twelve case studies also had the explicit aim of generating cost information for broader policy processes. In terms of pricing, two case studies reported on a costing exercise that was designed to inform capitation payment rates [16,31], six studies aimed at generating cost weights for DRGs [4,16,36]^1^ or unspecified case groups [16,27,29]^2^ and three papers reported on estimation of the cost of health benefit packages [16,25]^3^. A final case study reviewed the available cost evidence for informing price setting in the National Health Insurance Fund, Kenya [39].

**Table 1.**
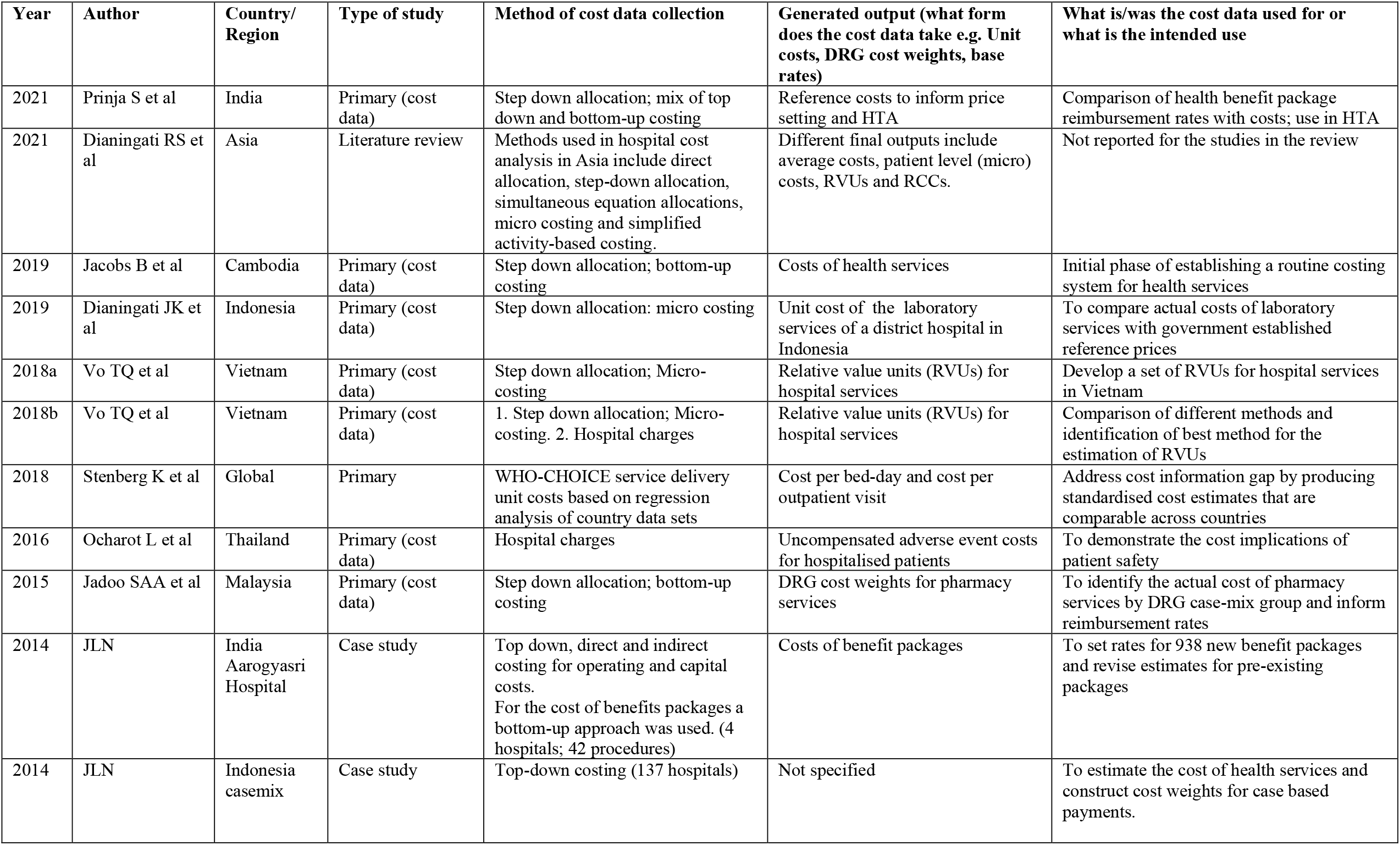
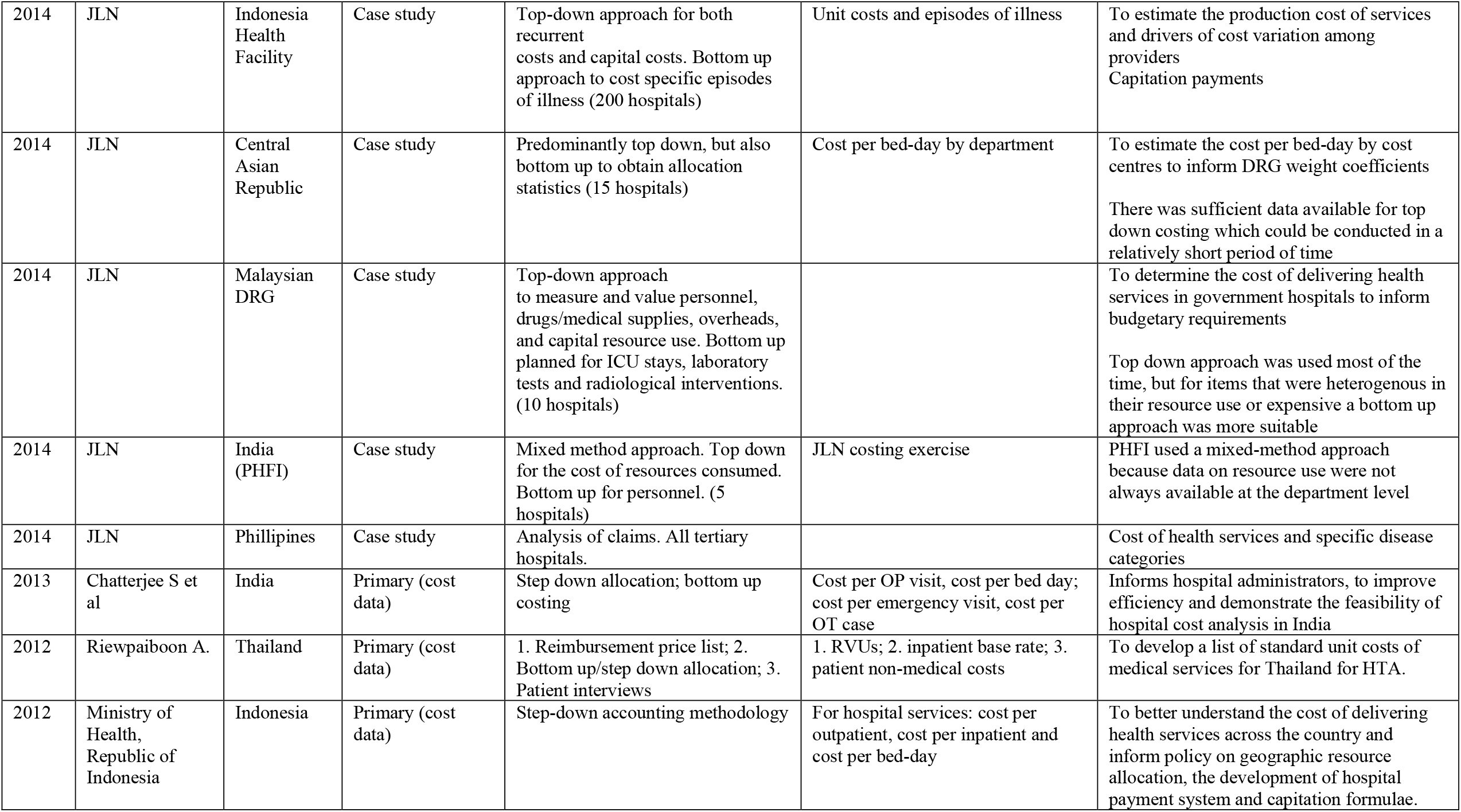
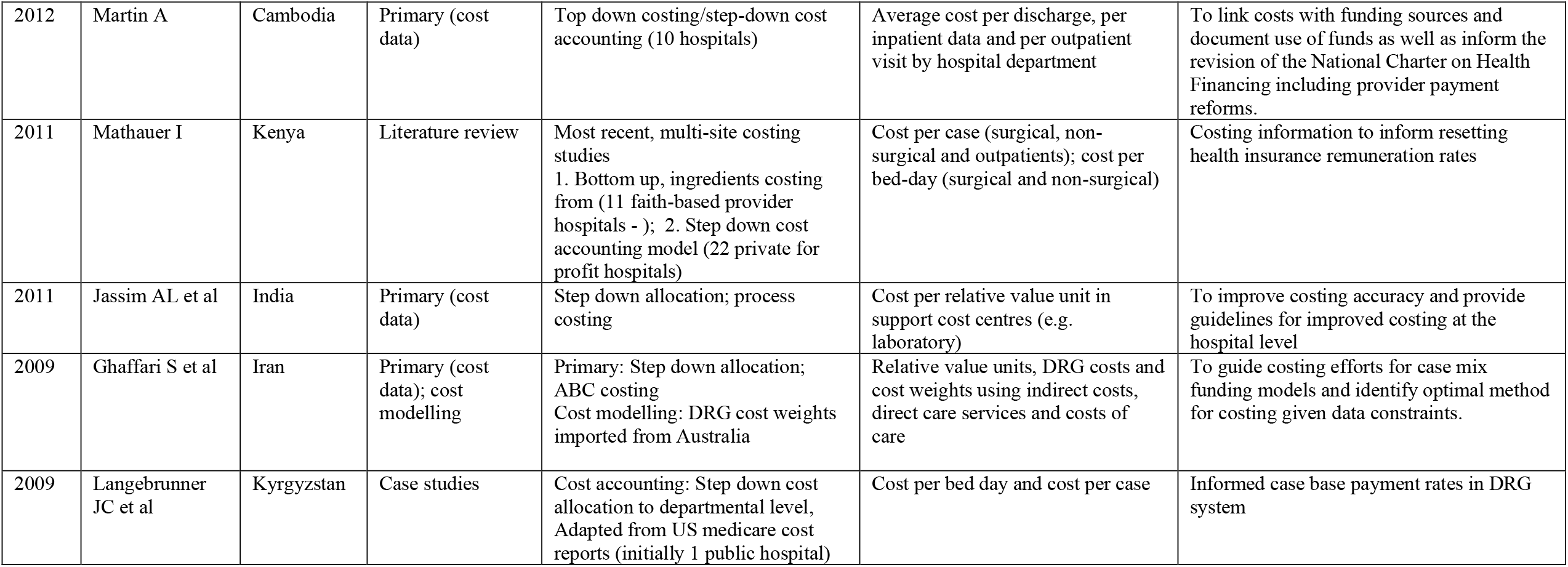
Costing evidence for tariff setting in hospital payment schemes in LMICs.

For the studies reporting costs, cost per service unit at the hospital level was the most frequently sited output e.g., cost per bed day, cost per admission and cost per outpatient visit. Three studies generated unit costs for specific services: cost per adverse event [26]; laboratory services [32]; and pharmacy services [36]. And a further three studies generated costs of health benefit packages [16,25]^3^. Relative value units were the primary output of 7 studies, one of which also explicitly estimated an inpatient base rate [27].

Fourteen of the case studies were commissioned by the local ministry of health or agency acting on their behalf. However, in many cases, it was not clear who had commissioned the costing or if the study was linked to the national policy process [14,16,23,24,26,29,30,36]^4^. Two studies evaluated different methods for generating robust relative value units [30,34].

### Papers reporting on how cost data informs the tariff-setting process

We identified 18 papers that provided explanation of the technical process of tariff-setting documenting experiences in 10 different countries (see Figure 3 and **Error! Reference source not found**.). Eight papers were published since the beginning of 2020, 8 papers in the period 2015-2019, 4 papers in the period 2010-2014, and 2 papers were published before 2010 (see Figure 4). The papers provided mixed levels of detail on the technical processes of price-setting and the strengths and weaknesses in each locality. The current tariff system, presence of an explanation of the price-setting process, the data used in price setting and resulting policy levers and implications are summarised in Figure 5.

**Figure 3.**
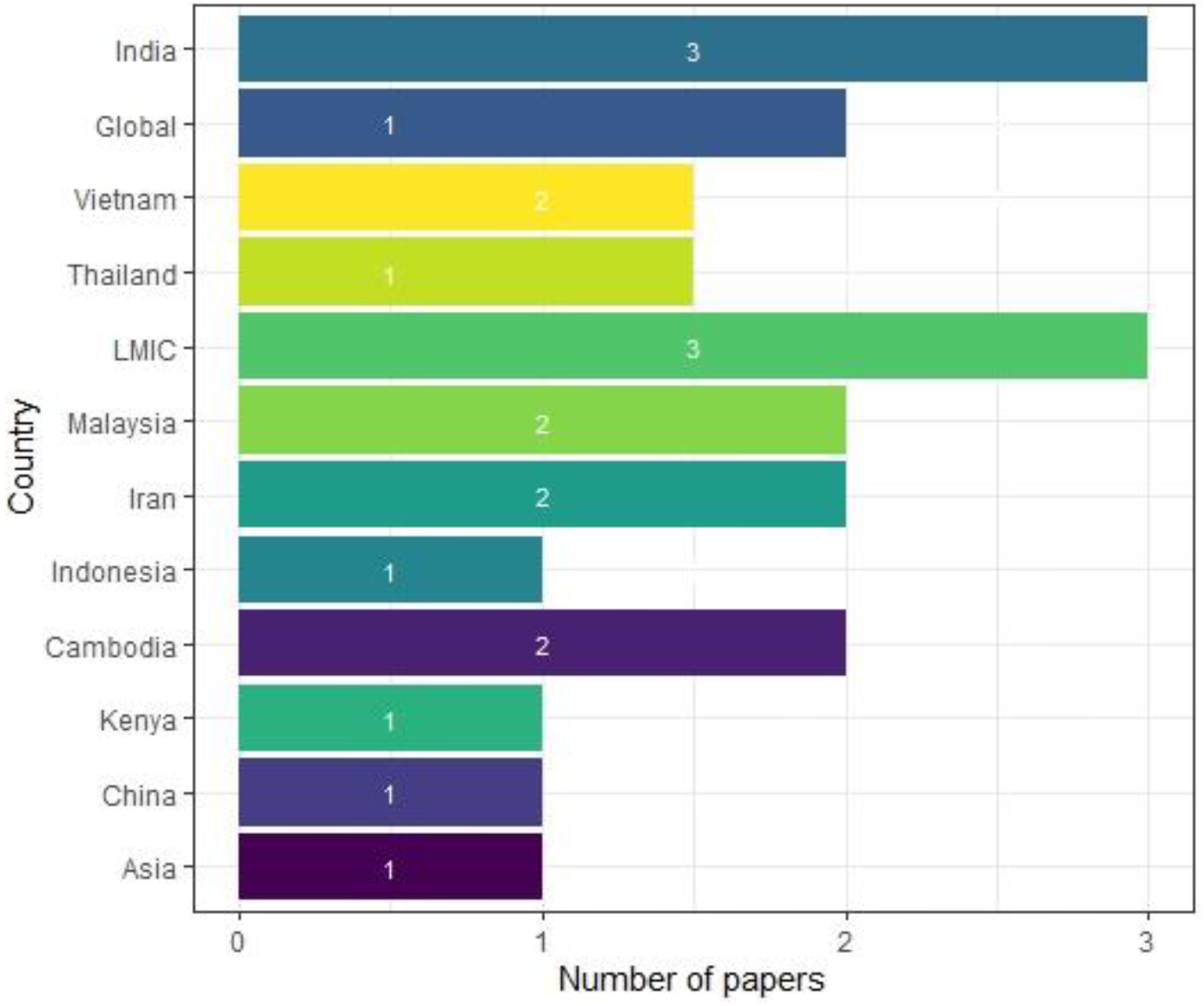
Number of papers explaining the tariff setting scheme by country.

**Figure 4.**
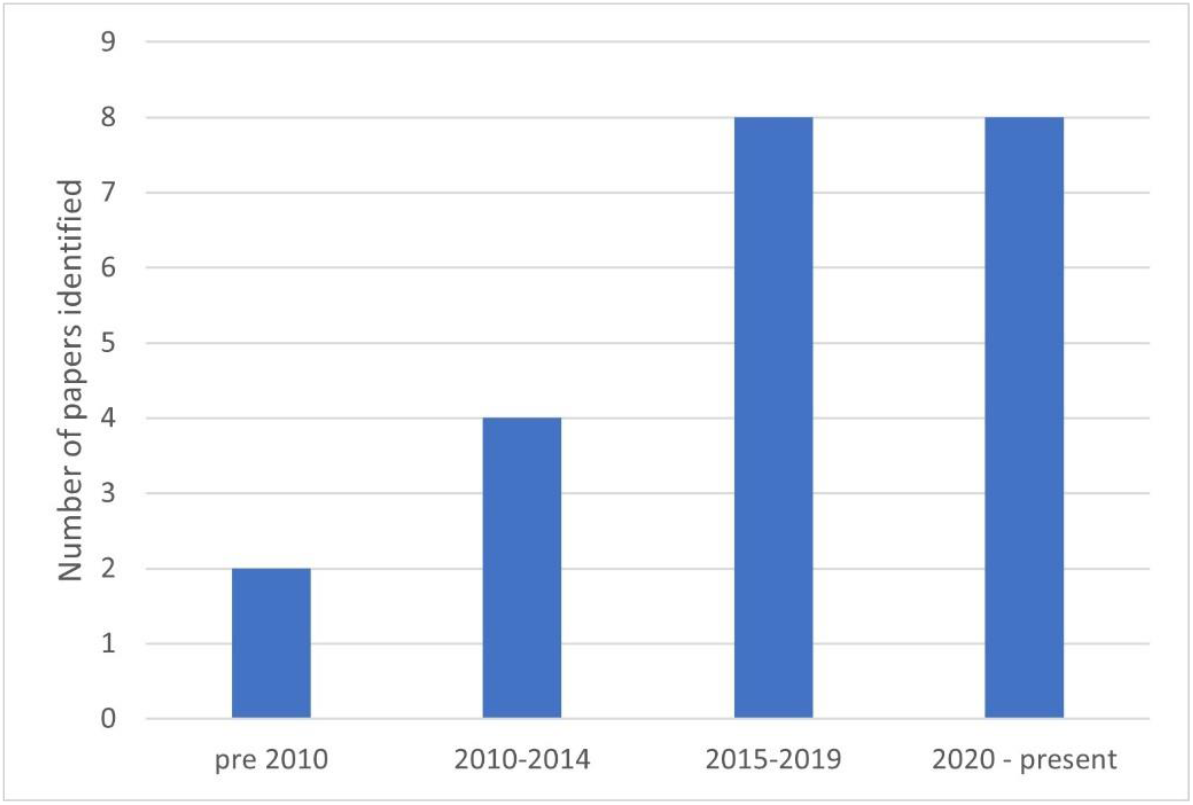
Number of papers by year of publication.

**Figure 5.**
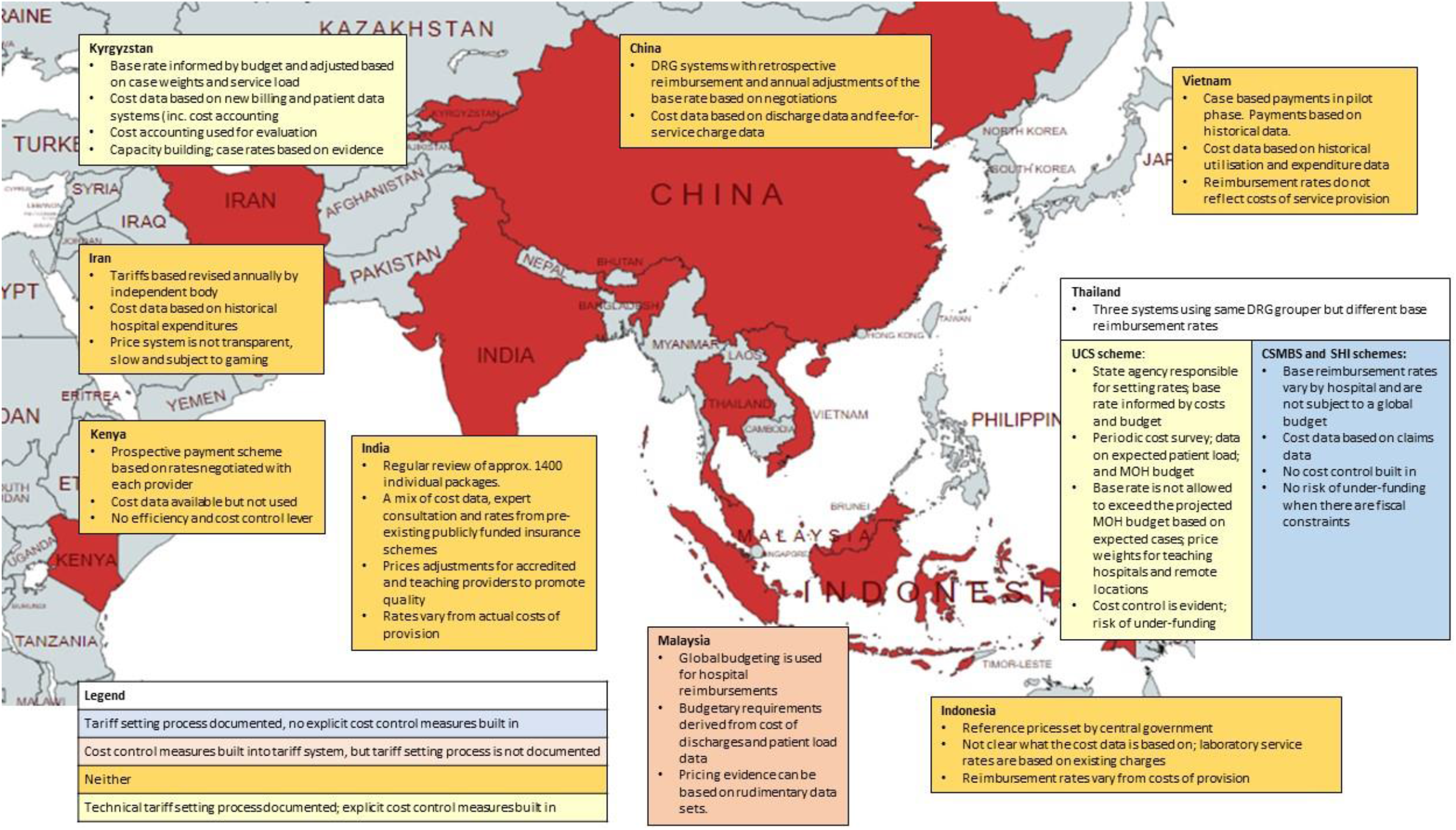
**Summary of evidence on the tariff setting process for case-based hospital payment in national health insurance schemes**

Only one study described tariff setting in Africa [39]. The paper reviewed the available evidence on costs for informing Kenya’s National Health Insurance Fund prices and was published in 2011. Although the cost information were considered reliable by all stakeholders, in part due to their involvement in the costing exercises, the costs had not been used for setting prices at the time of the study.

The other countries covered were all in Asia. In the Central Asia region, three papers focus on the reform of the tariff setting system in Kyrgyzstan [4,15,18]. The technical process of price setting is clearly documented. This process includes cost control measures derived from linking the reimbursement rates to the MOH budget. The Thai Universal Coverage Scheme (UCS) also provides an example in which cost control is built into the base rate through linkages to the budget. However, as figure 5 states, in Thailand, there are 3 government funded and implemented schemes. Although all Thai schemes use the same Thai-DRG grouper, the Civil Service Medical Benefit Scheme (CSMBS) and Social Health Insurance (SHI) schemes do not use cost control mechanisms, as the rates are not linked to an overall budget and are different rates for different hospitals [1,6,18,27].

The Kyrgyz reforms were found to be vulnerable to gaming as the system does not make full use of the data available potentially leading to misclassification of diagnoses. This potential for gaming is also highlighted in Iran. In contrast to the systematic introduction and use of cost accounting in Kyrgyzstan, Iran’s price setting process involves a technical assessment by an independent body but with limited transparency (see Figure 5) [33,34]. Doshmangir et al note that without an objective and explicit mechanism in the updating of medical tariff and no structure to effectively manage conflicts of interest, the pricing system has in effect become “a tool for revenue manipulation” [33].

Challenges also arise when reimbursement rates are not based on cost evidence. In India’s national insurance programme for the poor and vulnerable, the government used existing information to set reimbursement rates while establishing a review system to allow for modification and improvement over time (see Figure 5) [15]. However, the method in which information from costing studies, experts and rates under previous schemes is compiled is not transparently reported on. Studies show that the rates vary considerably from actual costs (42% of HBPs had a price less than 50% of the true cost in 2018) [25]. This could affect the recruitment of providers, the coverage and quality of care, and bring the rates themselves into doubt [41]. Indonesia faces a similar problem in respect of laboratory services. Dianingati et al, report on a lack of transparency in the development of the reference prices set by the government and that the true costs of service delivery are 40-53% higher than the reference price. However, validation of the rates is difficult as data on the cost of healthcare services is still limited to a few services, in focal geographical areas, restricted to the public sector with few published and readily accessible cost data analyses/ data sets [20,25].

Both the Thai UCS and Kyrgyz price-setting systems use cost accounting to inform their base rates and case weights. Langebrunner et al note how cost accounting has been used as an evaluation tool and allowed for tariff adjustments based on evidence so that payments match services in Kyrgyzstan. Similarly, in the Thai UCS scheme, a key feature of the tariff setting is the cost information on which pricing for the UCS is based. This is collected on a periodic basis in a cost survey and has evolved from initial work using an RVU method and “top level” hospital cost data [27] to a 900 hospital survey. While no study described how the system has reformed, the papers note that the gradual, step wise implementation allowed for institutional and technical capacity building.

There was less detail reported on the tariff setting process in China, Indonesia, Malaysia and Vietnam. The one study documenting tariff setting processes in China raises concerns that the schemes used retrospective payment system and fails to build in efficiency and cost control [18]. In Vietnam there was also concern that the fees and the payment schemes bore little relationship to the costs of delivering services although the relative value unit method used to calculate rates for the capitation scheme was relatively simple. Similarly, while the Malaysian system was designed for global budgeting, it also demonstrates that pricing evidence can be based on skeletal data sets such as those that focus on large expenditure items and patient data that are feasible to collect [1].

## DISCUSSION

### Cost systems increase the policy evidence base

Cost systems can create a transparent evidenced based process for price-setting and these systems need to generate good quality data, based on accepted methodologies. Studies from Iran and Thailand emphasise how important the cost system is in the setting of health benefit package/ DRG prices, to minimise gaming and prevent cost escalation [6,33]. Leaving base rates open to negotiation at the individual provider level with minimal evidence on costs and efficiency of service provision, leaves the system vulnerable to gaming.

### Cost evidence can increase efficiency of service provision

Creating a tariff setting system that does not use costs based on empirical evidence can embed inefficiencies and possibly make it more difficult to implement costing in the future [33]. A centralised cost accounting system, such as was developed in Kyrgyzstan, was considered a major strength of the broader health system reforms – allowing for policy reform to anticipate expenditure needs and enabling the government to effect change more effectively.

### Some data is better than no data but its important to plan early for sustainable institutionalized cost systems

On the other hand, it is also recognised that such exercises are expensive, and a persistent issue is that of overcoming the capacity constraints to generate this information. While obtaining good quality cost information can be an important aid to evidence-based decision-making, it is important to be aware of the trade off in accuracy and resources needed to generate the information. Where resources are highly constrained, any data can be better than no data, particularly if the data are reported transparently and how the data informs decisions is clearly communicated and accounted for. If cost accounting is not the norm and the budget is limited, costing for price setting may need to start with simpler methods, using, for example, expenditure data, relative value units and smaller samples of facilities. Alternative approaches, for example in India, Cambodia and Kenya, have started with the implementation of baseline multi-site costing studies. Although these are one off exercises, and therefore cannot identify drivers of efficiency, they provide an evidence base and good practice on which to build. The costing itself can be a way to bring stakeholders into the price setting process and build capacity for future costings.

The example of Kyrgyzstan shows how implementing a cost system is a slow, gradual and complex process. The established costing systems identified in the literature illustrate how a cost system has evolved from one-off exercises and developed into complex system with increasing numbers of participating providers (Thailand, Kyrgyzstan, China).

### Costing needs and methods vary and need to be appropriate for the setting

Methods for cost data collection also need to be appropriate for the setting. Studies from Thailand and Vietnam compare different approaches to obtaining the base rate and cost weights for health technology assessment and pricing. They compare micro costing with relative value unit approaches and find both to be feasible with micro-costing being highly resource intensive. The costing methods tend to follow the same principles using top-down allocation methods supplemented with bottom-up costing if resources allow, for some specific inputs. In Malaysia, one study demonstrated the feasibility of using the electronic prescribing system to generate DRG weights, although it was recognised that these were not available in most facilities.

### A transparent set of principles for translating cost evidence into base rates and weights is needed

As well as cost data collection, a systematic method for translating costs into prices or reimbursement rates helps avoid skewed incentives within the prices, evident in the unexplained differences between costs and reimbursement rates found in India and Indonesia [25,32]. Langenbrunner’s reporting of the Kyrgyzstan case study provides the most comprehensive description for the calculation of the base rate and case-based weightings and how to use these to set reimbursement rates [4]. Patharanarumol et al also describe the principles applied for estimating the base rates and weights in the Thai UCS scheme [6]. For both settings, explicitly accounting for the budget in the price estimation using an “economic adjustment” is a key mechanism of cost control. This level of transparency is not apparent elsewhere in the literature identified. For example, while the different strands of information used for price setting are documented in the reports on India, the method for combining this information is not reported [25,42]. And, in Iran, the lack of such a methodology was reported as a significant problem for the DRG system as a whole leading to price manipulation by different stakeholders [33].

### Limitations

The scoping review has found a very limited level of evidence around how cost and price evidence is used in the price-setting process at the country level. In addition, many of the studies are old and may be outdated. While this can be in part due to the infancy of many of the price-setting systems and reforms, the nature of the review may have limited the evidence generated. While early reforms might be reported for some countries, it was not possible to determine how the tariff-setting processes have evolved and are currently being implemented. For example, Vietnam’s pilot study was published in 2014 but there were no corresponding papers documenting next steps; nor did we identify more recent reports on Kenya and Cambodia where costing evidence from large multi-site studies were identified. Further, the terminology required to search for the role of cost evidence in price-setting in the literature is poorly defined. The terms cost and price are used in many different ways to mean different things which may have led to some omissions. In addition, during the screening process many articles were identified on the process of developing DRG type reforms, but few focussed on the price-setting process and how cost evidence is used in the price-setting process. To address this we extended the search and performed additional searches using the key words identified in the initial papers found that met the inclusion criteria. Our review of the grey literature was limited to a google search and snowballing from references that were identified in the initial search. It is likely evidence in this area lies in government and donor reports and we may have missed these. Restricting reports to the English language may have compounded this.

Despite these limitations and the few country case studies identified, consistent themes were evident around the need to use cost information using a systematic methodology, reporting this transparently and working with providers to develop the system. Our review confirms that cost evidence can increase efficiency of service provision by increasing the policy evidence base. To generate this evidence, countries need to build cost systems appropriate to the setting and data availability but allowing for and investing in increasing complexity as data systems improve. Even while national costing surveillance should be an aspiration, prices should be set using cost evidence which can take the form of one off costing studies, or even hospital charges. Especially in the absence of national surveillance, the method in which these data are then used to set base rates and price weights should be part of a transparent process that involves all stakeholders and takes account of heterogeneity in costs driven by demand side (e.g. condition or patient specific) and supply side (e.g. hospital location) factors.

## CONCLUSION

LMICs are increasingly turning to insurance-based models of healthcare and private sector providers to increase coverage of the poor and vulnerable. To help achieve value for money within these universal health coverage goals, publicly financed insurance schemes need to account for budget constraints, encourage efficient health service delivery and use good quality evidence transparently in setting reimbursement rates. Documentation of the good practice and the challenges of generating cost evidence and creating costing systems for informing reimbursement decisions in resource poor settings are lacking. While there are accepted and widely used methods for generating cost information, countries need to build more sustainable cost systems and adopt more transparent systems and methodologies for translating costs into prices.

## Data Availability

All data produced in the present study are available upon reasonable request to the authors

## ACKNOWLEDGEMENTS

Grateful thanks to Yi-Ling Chi and Lydia Regan at Center for Global Development for valuable comments and advice to the search strategy.

## AUTHORS’ CONTRIBUTIONS

- Concept and design: LG, HAS, SG, AM
- Acquisition of data: LG, SG
- Analysis and interpretation of data: LG, SG
- Drafting of the manuscript: HAS, LG, SG
- Critical revision of paper for important intellectual content: HAS, LG, SG
- Obtaining funding: LG, AM
- Supervision: LG, AM

## Footnotes

1 Joint Learning Network case studies: Central Asian Republics

2 Joint Learning Network case studies: Indonesia Ministry of Health

3 Joint Learning Network case studies: PhilHealth and India Aaorgyasri

4 Joint Learning Network case studies: India – Public Health Foundation of India

## REFERENCES

1 Barber SL, Lorenzoni L, Ong P. Price Setting and Price Regulation in Health Care. OECD 2019. doi:10.1787/ed3c16ff-en

2 Tan SY, Melendez-Torres GJ. Do prospective payment systems (PPSs) lead to desirable providers’ incentives and patients’ outcomes? A systematic review of evidence from developing countries. Health Policy and Planning 2018;33:137–53. doi:10.1093/HEAPOL/CZX151

3 Robyn PJ, Sauerborn R, Bärnighausen T. Provider payment in community-based health insurance schemes in developing countries: a systematic review. Health Policy and Planning 2013;28:111–22. doi:10.1093/heapol/czs034

4 Langenbrunner JC, Cashin C, O’Dougherty S. Designing and Implementing Health Care Provider Payment Systems : How-To Manuals. 2009.

5 Government of Australia. Australian Public Hospitals Cost report. 2016.https://www.ihpa.gov.au/publications/australian-public-hospitals-cost-report-2013-2014-round-18

6 Patcharanarumol, W Panichkriangkrai, W Sommanuttaweechai, A Hanson K, Wanwong, Y Tangcharoensathien V. Strategic purchasing and health system efficiency: A comparison of two financing schemes in Thailand. PLoS ONE 2018;13.https://pubmed.ncbi.nlm.nih.gov/29608610/ (accessed 20 Jan 2021).

7 Raulinajtys-Grzybek M. Cost accounting models used for price-setting of health services: An international review. Health Policy 2014;118:341–53. doi:10.1016/j.healthpol.2014.07.007

8 Bredenkamp C, Bales S, Kahur K. Transition to Diagnosis-Related Group (DRG) Payments for Health Lessons from Case Studies. Washington DC: 2020.

9 Barber SL, Lorenzoni L, Ong P. Price Setting and Price Regulation in Health Care. OECD 2019. doi:10.1787/ed3c16ff-en

10 Carroll N, Lord JC. The Growing Importance of Cost Accounting for Hospitals. J Health Care Finance 2016;43:172./pmc/articles/PMC6910125/ (accessed 7 Jul 2021).

11 García-Cornejo B, Pérez-Méndez JA. Influence of cost systems on efficiency. An analysis of Spanish hospitals using public national databases. Revista de Contabilidad-Spanish Accounting Review 2020;23:249–62. doi:10.6018/rcsar.365031

12 Barber SL, Lorenzoni L, Ong P. Price Setting and Price Regulation in Health Care. OECD 2019. doi:10.1787/ed3c16ff-en

13 Munn Z, Peters MDJ, Stern C, et al. Systematic review or scoping review? Guidance for authors when choosing between a systematic or scoping review approach. BMC Medical Research Methodology 2018;18:1–7. doi:10.1186/S12874-018-0611-X/TABLES/1

14 Stenberg K, Lauer JA, Gkountouras G, et al. Econometric estimation of WHO-CHOICE country-specific costs for inpatient and outpatient health service delivery. Cost Effectiveness and Resource Allocation 2018;16:11. doi:10.1186/s12962-018-0095-x

15 Barber SL, O’Dougherty S, Vinyals Torres L, et al. Other considerations than: how much will universal health coverage cost? Bull World Health Organ 2020;98. doi:10.2471/BLT.19.238915

16 Joint Learning Network. Costing of Health Services for Provider Payment: A Practical Manual. 2014.

17 Mathauer I, Wittenbecher F. Hospital payment systems based on diagnosis-related groups: experiences in low-and middle-income countries. Bull World Health Organ 2013;91:746–756A. doi:10.2471/BLT.12.115931

18 Bredenkamp C, Bales S, Kahur K. Transition to Diagnosis-Related Group (DRG) Payments for Health Lessons from Case Studies. 2020.

19 Dianingati RS, Riewpaiboon A. Unit cost analysis of medical service in asia: A systematic review. doi:10.29090/psa.2021.02.19.034

20 Prinja S, Chauhan AS, Rajsekhar K, et al. Addressing the Cost Data Gap for Universal Healthcare Coverage in India: A Call to Action. Value in Health Regional Issues 2020;21:226–9. doi:10.1016/J.VHRI.2019.11.003

21 National Health Authority. Journey from HBP 1.0 to HBP 2.0. 2019.

22 KPMG. Ayushman Bharat — A big leap towards Universal Health Coverage in India. 2019.

23 Chatterjee S, Levin C, Laxminarayan R. Unit Cost of Medical Services at Different Hospitals in India. PLoS ONE 2013;8:e69728. doi:10.1371/journal.pone.0069728

24 Jassim AL. Cost Measuring of Medical Services in a Hospital using Relative Value Units (RVUs) – based Process Costing System. undefined 2011.

25 Prinja S, Singh MP, Rajsekar K, et al. Translating Research to Policy: Setting Provider Payment Rates for Strategic Purchasing under India’s National Publicly Financed Health Insurance Scheme. Applied Health Economics and Health Policy 2021;:1–18. doi:10.1007/s40258-020-00631-3

26 Ocharot L, ‘Sriratanaban J’, Ngamkiatphaisan. Sureerat’ ‘, et al. Estimating uncompensated medical care cost as a result of adverse events in a university hospital in Thailand. Asian Biomedicine 2016;10:261–7.

27 Riewpaiboon A. Standard cost list for economic evaluation in Thailand. Value in Health 2012;15:A645–A645. http://www.embase.com/search/results?subaction=viewrecord&from=export&id=L70915622 %5Cn http://dx.doi.org/10.1016/j.jval.2012.08.255

28 Minh H van, Phuong NK, Özaltın A, et al. Costing of commune health station visits for provider payment reform in Vietnam. Global Public Health 2015;10. doi:10.1080/17441692.2014.944929

29 Vo TQ, Chaikledkaew U, Minh H, et al. Development of Relative Value Units for Unit Cost analysis of Medical Services in Vietnam. Asian Journal of Pharmaceutics 2018;12:S19–26.

30 Vo TQ, Chaikledkaew U, Minh H, et al. Hospital cost analysis in developing countries: A methodological comparison in Vietnam. Asian Journal of Pharmaceutics 2018;12:S8–18.

31 Ministry of Health Republic of Indonesia. THE COSTS OF DELIVERING HEALTH SERVICES IN INDONESIA: REPORT ON A PROSPECTIVE SURVEY 2010-2011. 2012.

32 Dianingati RS, Riewpaiboon A, Youngkong S. Indonesia Hospital Cost Analysis: a Micro-Costing Approach. Jurnal Kesehatan Masyarakat 2019;14. doi:10.15294/kemas.v14i3.15627

33 Doshmangir L, Rashidian A, Kouhi F, et al. Setting health care services tariffs in Iran: half a century quest for a window of opportunity. International Journal for Equity in Health 2020;19. doi:10.1186/s12939-020-01224-1

34 Ghaffari S, Doran C, Wilson A, et al. Investigating DRG cost weights for hospitals in middle income countries. The International Journal of Health Planning and Management 2009;24:251–64. doi:10.1002/hpm.948

35 Rasiah D. Why Activity Based Costing (ABC) is still tagging behind the traditional costing in Malaysia? Journal of Applied Finance & Banking 2011;:83–106.

36 Ali Jadoo SA, Aljunid SM, Nur AM, et al. Development of MY-DRG casemix pharmacy service weights in UKM Medical Centre in Malaysia. DARU Journal of Pharmaceutical Sciences 2015;23. doi:10.1186/s40199-014-0075-4

37 Martin A. Cambodia Hospital Costing and Financial Management Study. 2012.

38 Jacobs B, Hui K, Lo V, et al. Costing for universal health coverage: insight into essential economic data from three provinces in Cambodia. Health Economics Review 2019;9:29. doi:10.1186/s13561-019-0246-6

39 Mathauer I. Setting health insurance remuneration rates of private providers in Kenya: the role of costing, challenges and implications. The International Journal of Health Planning and Management 2011;26. doi:10.1002/hpm.1038

40 Zhao C, Wang C, Shen C, et al. Diagnosis-related group (DRG)-based case-mix funding system, a promising alternative for fee for service payment in China. BioScience Trends 2018;12. doi:10.5582/bst.2017.01289

41 Prinja S, Bahuguna P, Singh MP, et al. Determining Price Weights for Differential Case-Based Payments under India’s National Publicly Financed Health Insurance Program. Under review: 2022.

42 National Health Authority. Journey from HBP1.0 to HBP2.0. New Delhi: 2019.

