## Supplement for "The role of healthcare cost accounting in pricing and reimbursement in low- and middle-income countries"

### Supplementary materials

#### Search strategy

| Database | Search terms |
| --- | --- |
| **Web of Science** | (ALL=(("case*mix" or "cost systems" or "cost*accounting" or "ref*costs" or "resource weights" or "cost*weights" or "national reimbursement" or "DRG" or "hospital payment systems" or "fee*for*service"))) AND ALL=("LMIC" OR "low resource settings" OR "developing countries") |
| **Econlit** | ("case*mix" or "cost systems" or "cost*accounting" or "ref*costs" or "resource weights" or "cost*weights" or "national reimbursement" or "DRG" or "hospital payment systems" or "fee*for*service").mp. [mp=heading words, abstract, title, country as subject]<br><br>No hits when combined with lmics |
| **Medline** | 1. ("case mix" or "DRG" or "case based payment" or "hospital payment system" or "reimbursement" or "resource value unit" or "cost systems" or "cost accounting" or "reference costs" or "resource weights" or "cost weights" or "Price setting" or "service weights")<br>2. "Costs and Cost Analysis"/ or National Health Programs/ or diagnosis-related groups/ or hospitalization/<br>3. 1 OR 2<br>4. Developing Countries/<br>5. "universal health*<br>6. Universal Health Insurance/ or "Delivery of Health Care"/ or Insurance, Health/<br>7. 5 OR 6<br>8. 3 AND 4 AND 7 |
| **PubMed** | ("reference cost*" OR "reference price*") AND health* AND national* AND list*)<br>Since 2000 |

#### List of studies identified and included in the review

| Author | Year | Country | Aim of paper in relation to costing (rationale for inclusion) | Type of study | Cost data reported | Comments on country's cost system |
|---|---|---|---|---|---|---|
| Langebrunner JC et al | 2009 | LMIC (Kazakhstan, Kyrgyzstan) | Manual on methods for setting up provider based systems; provides methods and case study of costing accounting for case based payments | Case studies | Yes | Yes |
| Joint Learning Network | 2014 | LMIC | Costing specific resource for lmics | Case studies | Yes | Yes |
| Martin A | 2012 | Cambodia | Costing in Cambodian hospital | Primary (cost data) | Yes | Yes |
| Ministry of Health, Republic of Indonesia | 2012 | Indonesia | Report on a costing study for Indonesia | Primary (cost data) | Yes | Yes |
| Ghaffari S et al | 2009 | Iran | Costing for DRGs in Iran | Primary (cost data) | Yes | Yes |
| Mathauer I | 2011 | Kenya | Role of costing in setting insurance reimbursement rates in Kenya | Literature review | Yes | Yes |
| Jadoo SAA et al | 2015 | Malaysia | Documenting the development of DRG cost weights in pharmacy in Malaysia | Primary (cost data) | Yes | Yes |
| Dianingati JK et al | 2019 | Indonesia | Single site cost data collection to inform price setting | Primary (cost data) | Yes | Yes |
| Jacobs B et al | 2019 | Cambodia | Multiple site cost data collection to inform national policy including reimbursement rates | Primary (cost data) | Yes | Yes |
| Ocharot L et al | 2016 | Thailand | Cost implications of adverse events in DRG system | Primary (cost data) | Yes | Yes |
| Prinja S et al | 2021 | India | Comparing cost data with reimbursement rates in ABPMJAY, India | Primary (cost data) | Yes | Yes |
| Riewpaiboon, A. et al | 2012 | Thailand | Development of Relative Value Units for Unit Cost analysis of Medical Services in Thailand | Primary (cost data) | Yes | Yes |
| Vo TQ et al | 2018a | Vietnam | Development of Relative Value Units for Unit Cost analysis of Medical Services in Vietnam | Primary (cost data) | Yes | Yes |
| Vo TQ et al | 2018b | Vietnam | Comparison of hospital costing methods in Vietnam | Primary (cost data) | Yes | Yes |
| Dianingati RS et al | 2021 | Asia | Literature review of medical service costs in Asia | Literature review | Yes | No |
| Jassim AL et al | 2011 | India | Testing for RVU method for costing in hospitals in India | Primary (cost data) | Yes | No |
| Chatterjee S et al | 2013 | India | Hospital costing study | Primary (cost data) | Yes | No |
| Stenberg K et al | 2018 | Global | Estimating unit costs of health services at a country level based on global dataset | Primary (cost data) | Yes | - |
| Lian LL et al | 2014 | Taiwan | Assessing incentives in DRGs | | No | Yes |
| Barber S et al | 2019 | Global (India, Malaysia, Thailand) | Manual and case studies on price setting for case-based payment | Case studies | No | Yes |
| Bredenkamp C et al | 2020 | Global (China, Kyrgyz Republic, Thailand) | Case studies of DRG transitions | Case studies | No | Yes |

| Author | Year | Country | Aim of paper in relation to costing (rationale for inclusion) | Type of study | Cost data reported | Comments on country's cost system |
|---|---|---|---|---|---|---|
| Zhao C et al | 2018 | China | Document China's experiences with shifting to case based payment schemes | Case studies | No | Yes |
| Prinja S et al | 2020 | India | Commentary on cost data for policy | Commentary | No | Yes |
| Mathauer I et al | 2013 | LMIC | Literature review of DRG experiences in LMICs | Literature review | No | Yes |
| Doshmangir L et al | 2020 | Iran | To document Iran's experience of tariff setting | Primary | No | Yes |
| Hoang VM, et al | 2014 | Vietnam | Reporting on costing in Vietnam for provider payment reform | Primary | No | Yes |
| National Health Authority | 2019 | India | Describes process of updating HBP package rates | Primary | No | Yes |
| Patcharanarumol K et al | 2018 | Thailand | Comparing strategic purchasing in two financing schemes | Primary | No | Yes |
| Rasiah, D et al | 2011 | Malaysia | Comparing methods for costing of health services in Malaysia | Primary | No | Yes |
| KPMG | 2019 | India | Overview of AB-PMJAY reform and financing mechanisms | Report | No | Yes |
| Barber S et al | 2020 | Global | To provide policy recommendations on estimating the cost of UHC | Case studies | No | Yes |
| Wagstaff A et al | 2007 | East Asia | Lessons learned in Asia for financing reforms including provider payment | Primary | No | Yes |
| Hu S et al | 2008 | China | Commentary | Commentary | No | No |
| Zeng W et al | 2018 | Global | Examining role of PBF in strengthening health systems | Commentary | No | No |
| Beck E et al | 2012 | LMIC | Describes the financial information required by policy makers and other stakeholders to enable them to make evidence-informed decisions and reviews the quantity and quality of the financial information available, | Literature review | No | No |
| Zou K et al | 2020 | China | Literature review of impact of case based payments in China | Literature review | No | No |
| Bertram M et al | 2017 | Global | Estimating unit costs for disease control programmes | Primary | No | No |
| Immunisation Costing Action Network | 2018 | LMIC | Costing of immunisation programmes | Primary | No | No |
| Jian W et al | 2016 | China | Assessing capacity of information system to implement DRGs (not cost system) | Primary | No | No |
| Watkins D et al | 2020 | LMIC | Resource requirement estimation of model health benefit packages (essential services) | Primary | No | No |
